# A co-designed website (TeenFit) to promote physical activity in adolescents through mobile apps: a study protocol

**DOI:** 10.1101/2025.11.25.25340945

**Authors:** Anna Seiterö, Alicia Ruiz-Moreno, Adriana Alcarazo, Carmen M. Garrido-Gonzalez, María I. Ballesta-Rodríguez, Pontus Henriksson, Rodrigo A. Lima, Yolanda Álvarez-Pérez, Rafael E. Reigal, Henar Campos-Paíno, Juan Ángel Bellón, Emma Motrico, María Flores-López, María Rodriguez-Ayllon

**Author notes:** **Corresponding Authors**: Anna Seiterö; Department of Health, Medicine and Caring Sciences, Linköping University, SE-581 83 Linköping, Sweden, María Rodriguez-Ayllon; Biomedical Research Institute of Málaga and Nanomedicine Platform (IBIMA Plataforma BIONAND); C. Severo Ochoa, 35, Campanillas, 29590 Málaga, Spain.

## Abstract

**Introduction:** Adolescence is a critical period marked by physiological and psychological changes that influence long-term health. Establishing healthy behaviors, particularly regular physical activity, is essential during this stage; however, around 80% of young people do not meet the World Health Organization’s physical activity recommendations. Digital technologies offer promising opportunities to promote health among youth, yet most physical activity apps lack scientific validation and usability. Unlike psychology, which has validated repositories to guide users in selecting mental health apps, physical activity lacks validated repository platforms, especially for adolescents. The project has a twofold aim: (1) to co-design a website, named the TeenFit website, that facilitates the selection of physical activity apps tailored to each adolescent’s individual needs and preferences; and (2) to investigate the barriers and facilitators for the implementation of the TeenFit website in real-world settings.

**Methods and analysis:** Adolescents and other stakeholders (i.e., caregivers, educators, and health professionals) will be engaged in the development of the TeenFit website app finder through a participatory design approach that involves three workshops, encompassing both a development phase and an implementation phase. Approximately 10-12 participants per stakeholder group will take part in smaller working groups (i.e., workshops) to collaboratively contribute to the co-design of the TeenFit website. TeenFit will include mobile applications available in the Spanish market, developed in both English and Spanish. These applications are currently being identified through two ongoing systematic reviews. In addition to qualitative data gathered during the audio- and video-recorded workshops, quantitative data will be collected on participants’ co-design experiences and the acceptability of the TeenFit website. Data analysis will involve descriptive statistics, qualitative content analysis for describing participants’ perspectives on what aspects (i.e., content, features, and layout) are important for the design of the website, and reflexive thematic analysis to identify barriers and facilitators to implementation.

**Ethics and dissemination:** The study was approved by the Research Ethics Committee in the province of Málaga (Spain) (REC ref: SICEIA-2024-003172). Research findings will be disseminated primarily via national and international conferences and publication in peer-reviewed journals. Patient and public involvement will inform further dissemination activities.

**Strengths and limitations of this study:**

- The major strength of the study is that it draws on both qualitative and quantitative data to generate meaningful insights into the co-design and implementation of the TeenFit website, including participants’ co-design experiences, preferences, sociodemographic characteristics, the website’s acceptability, and the barriers and facilitators to its implementation.
- Limitations include the use of a convenience sampling approach, which may lead to uneven demographic representation. A transparent reporting of participant characteristics will nevertheless allow readers to assess whether our findings are transferable to other settings.
- Offering one-to-one online participation alongside in-person workshops may mitigate accessibility barriers, though it could affect the dynamics of group interaction compared with fully in-person sessions.

## INTRODUCTION

Physical inactivity has been recognized as a major public health concern for decades [1]. Despite advances in technology that allow for easier monitoring and promotion of physical activity, global inactivity rates continue to rise [2–4]. According to the World Health Organization, in 2022, nearly one-third of adults worldwide (31.3%) were insufficiently active, an increase from 23.4% in 2000 [4]. These patterns suggest that current public health strategies are insufficient, and innovative approaches are needed to reverse these trends and reduce the burden of lifestyle-related diseases.

Adolescence represents a critical period for establishing healthy behaviors that often persist into adulthood. During this stage, individuals begin to take on adult responsibilities and form habits that can affect long-term health outcomes [5–7]. The WHO recommends that adolescents engage in at least 60 minutes of moderate-to-vigorous physical activity daily, yet over 80% fail to meet these guidelines [8]. This widespread inactivity places young people at increased risk for obesity, mental health challenges, and early onset of lifestyle-related diseases. Furthermore, these statistics highlight the limitations of existing interventions and underscore the urgent need for strategies that effectively engage adolescents and provide them with the tools and motivation to adopt and maintain an active lifestyle.

Digital technology offers a promising avenue to address these challenges, particularly among adolescents, who are considered “digital natives.” Mobile applications provide accessible, cost-effective, and flexible platforms for promoting physical activity, enabling users to exercise anytime and anywhere [9, 10]. However, the sheer number of available apps creates challenges for young people, caregivers, and health professionals who need to identify options that are safe, evidence-based, and user-friendly. To address this gap, we are currently conducting a systematic search of the Google Play Store alongside a scoping review to identify physical activity apps suitable for adolescents. We are focusing on apps whose design is informed by scientific methods, whose effectiveness on health outcomes has been evaluated, and that have undergone implementation studies. Both review protocols have been published in the OSF open-access database [11, 12]. While these reviews provide valuable insights for the scientific community by identifying gaps and promoting collaboration between industry and research, it is also essential to develop tools that make this information accessible and easy to use. Such tools can guide adolescents toward evidence-based apps and support them in adopting a more active and healthier lifestyle.

In some fields, such as psychology, web-based repositories have been developed to guide both patients and clinicians in selecting mental health apps based on validated quality criteria. For instance, the Division of Digital Psychology created an evaluation system and an online library of mental health applications that includes expert assessments, filters, and detailed app descriptions to support informed decision-making [13]. Yet, no such platform currently exists in the field of physical activity, particularly for adolescents. Developing a similar system could be instrumental in addressing the needs identified in this project by helping users, caregivers, and professionals select high-quality, scientifically supported physical activity apps.

## AIMS AND OBJECTIVES

### Aims

The overall aim of this project is to develop a co-designed website, in collaboration with adolescents, caregivers, educators, and health professionals, aimed at supporting young people to promote physical activity using third-party mobile applications that have been pre-screened by an expert committee to ensure their quality.

### Objectives

The project has a twofold aim: (1) to co-design a website, named the TeenFit website, that facilitates the selection of physical activity apps tailored to adolescents’ needs; and (2) to investigate the barriers and facilitators for the implementation of the TeenFit website in real-world settings (e.g., high schools, sports clubs, and healthcare settings).

## METHODS AND ANALYSIS

### Study context

This study is part of a larger research project that aims not only to co-design and evaluate the implementation feasibility of the TeenFit website, but also to systematically identify physical activity apps for young people aged 10 to 24 years, as described above. Although the systematic reviews informing this project include a wide age range, the TeenFit co-design and implementation described in this article will specifically focus on adolescents aged 16 to 19 years and their caregivers. This focus is based on two main reasons. First, in Spain, adolescents within this age range are legally able to make their own health-related decisions. Second, this period represents a critical developmental stage in which young people begin making independent choices that can significantly influence their long-term health and well-being.

### Study design

The TeenFit website will be developed through a participatory co-design approach that tailors digital health services to users’ needs, priorities, preferences, and builds on their skills. This methodology has been shown to achieve better results in terms of adherence, usability, and implementation compared to more traditional design approaches that do not include citizen participation [14, 15]. In this project, the co-design process (**Figure 1**) consists of a development phase and an implementation phase, during which three workshop sessions will be held with four groups of stakeholders: (1) adolescents, (2) caregivers, (3) educators, and (4) health professionals. Each workshop will focus on exploring stakeholders’ needs and desires (**workshop 1**), experiences, priorities, and preferences (**workshop 2**), and conditions influencing the acceptability and usability of the developed website in routine practice (**workshop 3**) (**Supplemental Material 1)**. While all participants from workshop 1 will be invited to participate in workshop 2, the third workshop session will additionally include a sample of individuals who have not been involved in the previous phases in order to prevent bias and promote new perspectives. In addition, while the development phase primarily incorporates qualitative data on stakeholders’ needs, priorities, and preferences, the implementation phase will comprise both qualitative and quantitative data. In accordance with a convergent mixed-methods study design [16], results from the quantitative and qualitative datasets will be merged and interpreted holistically using a joint display, visually bridging the findings to enhance and expand insights from the integrated analysis [17]. Results will be reported following guidelines for qualitative studies [18] and mixed-method studies [19].

**Figure 1.**
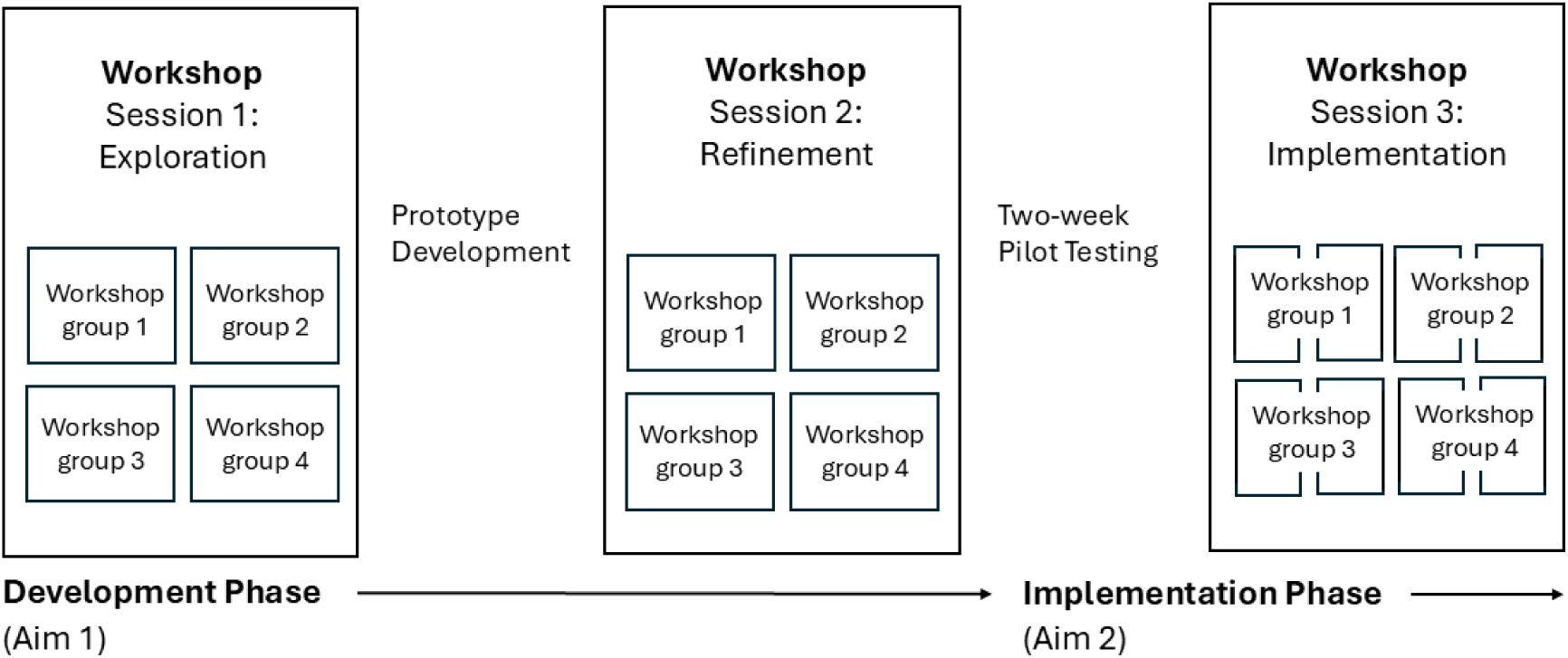
Overview of this two-phased participatory study design with four groups of stakeholders.

### Patient and public involvement

Inspired by Sanders and Stappers [20], we define co-design as the creativity of designers and people not trained in design working together in the design development process. In this approach, roles are diversified, and end users are positioned as experts of their experiences, taking a central role in generating knowledge, ideas, and concepts. Adolescents, caregivers, educators, and health professionals will therefore be actively involved both in the development and the implementation phases of this project. Stakeholder involvement provides valuable input on aspects that need to be considered to optimize the intervention and its potential for successful integration into key health-promoting settings targeting adolescents, such as high schools[21], sports clubs [22], and primary healthcare centers [23].

### Recruitment and Participants

Four groups of stakeholders will be recruited, with approximately 10 to 12 participants representing each group: adolescents aged 16-19 years; parents and other legal caregivers; educators (e.g., teachers and coaches); and health professionals (e.g., medical doctors, sports scientists, nurses, and physiotherapists). The total sample will comprise approximately 40–48 participants, a group size shown to provide sufficient feedback (i.e., information power) in previous focus-group [24] or co-creation studies [14, 25, 26].

Adolescents, caregivers, and educators will be recruited from secondary schools in the province of Málaga, Andalusia, Spain. Given the age range of 16 to 19 years, schools offering vocational training will be prioritized. The research team plans to visit secondary schools, present the project, and select one or two interested in collaborating. Secondary schools with cultural diversity and varying socioeconomic levels will be prioritized. A convenience sampling strategy will be used to recruit participants (adolescents, caregivers, and educators) within each school, using a Google Form to disseminate information about the project and provide an informed consent form. Efforts will also be made to ensure heterogeneity in terms of sociodemographic data (e.g., age, gender, socioeconomic status, or physical activity level). Health professionals will be recruited in the province of Jaén, Andalusia, Spain, leveraging previous collaborations and the willingness of local health centers to participate. The first step will involve a meeting with the district coordinator overseeing the eight primary care centers. For these groups, diversity in professional specialty will be prioritized.

Recruitment for the first workshop is scheduled to begin in November 2025, with the second workshop planned for May 2026 and the final one in June 2026. This timeline is approximate and may be adjusted based on recruitment challenges and participants’ preferences. If the target number of participants is not reached through schools and healthcare centers, additional recruitment strategies will include distributing flyers at high schools, universities, community centers, and relevant associations, as well as outreach via social media. No financial incentives will be offered; however, collaborating institutions will be acknowledged in any resulting publications to recognize their contributions.

### Workshops

Two workshop sessions will take place during the development phase, and a third workshop session will be conducted during the implementation phase, i.e., after a two-week pilot testing of the co-designed website (**Figure 1**). As shown in **Figure 2**, all workshops will have a similar structure that includes preparation, introduction, main activity, and evaluation (**Supplemental Material 1**), following the co-creation framework proposed by Álvarez-Pérez et al. [10] and Müssener et al. [27]. Preparation refers to activities that take place before participants attend, introduction refers to the formal procedures of outlining the agenda for the session, and evaluation refers to a quantitative assessment of participants’ experiences of the workshop session.

**Figure 2.**
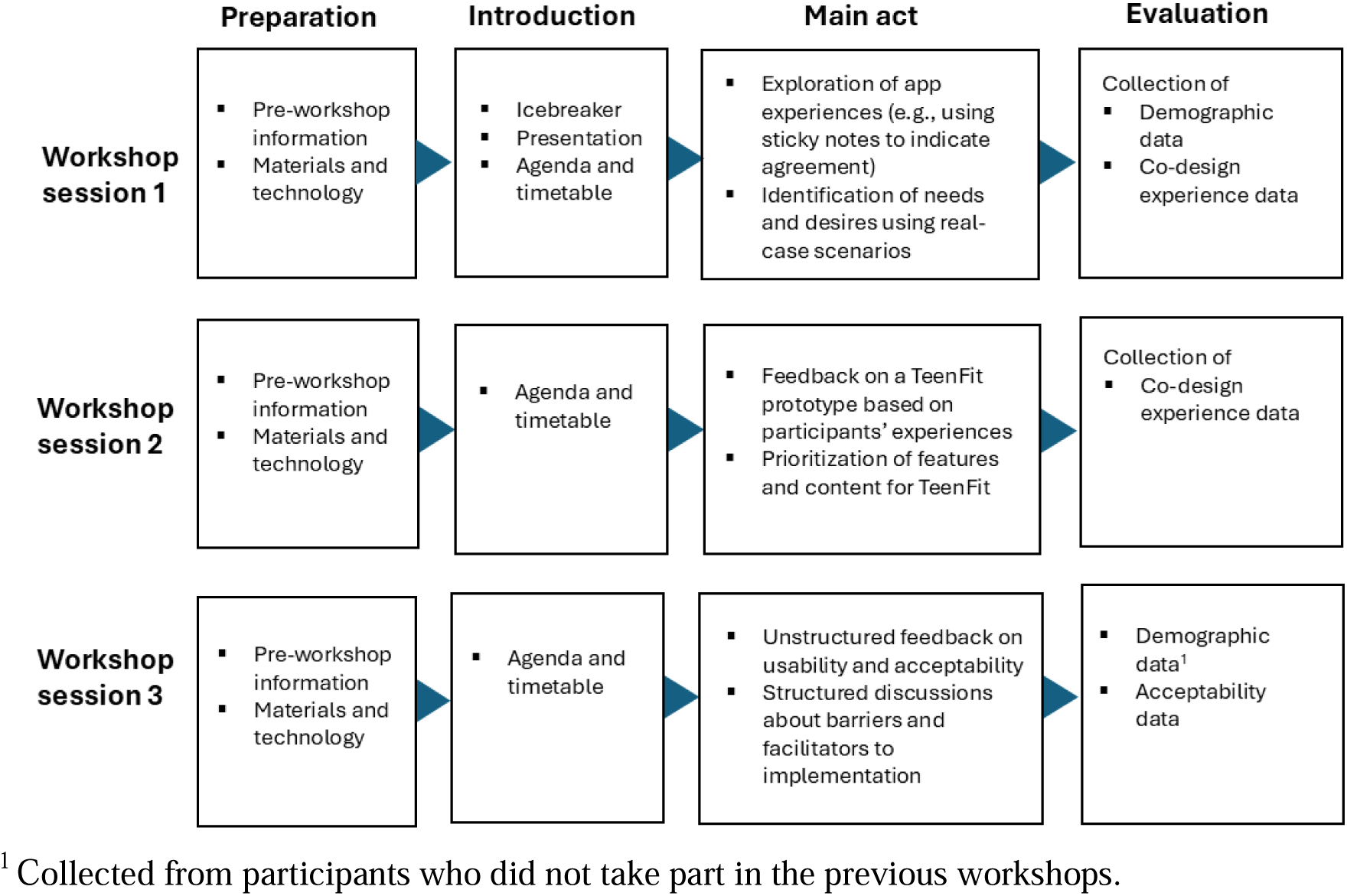
Script of the structure and content of the three workshop sessions within the TeenFit project.

The main activities include brainstorming exercises, scenario-based discussions, and prototype testing, allowing participants to provide feedback on design, functionality, and usability. To promote interaction and engagement, each stakeholder group will be divided into smaller subgroups of 5–6 participants while using digital and physical materials such as whiteboards, sticky notes, wireframe templates, colored cards, and interactive online tools. This structure allows for active participation, facilitates discussion, and helps participants feel valued and heard—factors that are known to enhance engagement in intervention design with youth populations [28]. Also, quantitative elements, such as color-coded voting or ranking exercises, will be jointly summarized and reviewed during the workshops to support transparent and collaborative decision-making.

More specifically, the main activity of the first workshop session will employ scenario-based brainstorming exercises to explore practical use cases. This type of exercise has been previously used in co-design interventions involving similar participants [29]. For example, in the group of health professionals, participants will discuss scenarios such as a healthcare provider recommending a physical activity app to an adolescent, considering factors like age, preferences, and goals (e.g., improving mental health). This approach aligns with best practices in co-design, where stakeholders collaboratively generate ideas to address real-world challenges [30].

The main activity of the second workshop session will involve presenting the first prototype of the TeenFit website to participants, followed by structured discussions to gather feedback on design elements, functionality, and overall user experience. Stickers will be used to prioritize key components of the intervention, while colored cards (green, yellow, red) will indicate the level of agreement with specific suggestions. This iterative process is crucial for refining digital health interventions and ensuring they meet the needs of the target audience. The workshops will follow a co-design protocol previously validated in similar digital health projects, such as the one described by Álvarez-Pérez et al., which has demonstrated effectiveness in engaging stakeholders and enhancing the usability and relevance of health interventions [14].

Finally, the main activity of the third workshop session will focus on participants’ experiences of using the website, and their perspectives regarding potential barriers and facilitators for implementing TeenFit in real-world settings (e.g., high schools, sports clubs, or healthcare settings). This means that the third workshop will take place after two weeks of pilot testing. Compared with the first two initial workshops, the third workshop session will be less practically oriented and, by its nature, be more of a focus group session structured around themes to discuss and prompts to stimulate discussion [31]. Also, for the third workshop, half of the participants will be drawn from the original co-design workshops, while the other half will be new participants who have not previously been involved. This strategy is intended to help minimize potential biases. Specifically, selection bias may occur if participants from earlier co-creation sessions differ systematically from others in terms of motivation or familiarity with TeenFit. Performance bias may arise if prior involvement influences how participants engage with workshop activities. Similarly, information bias may occur if previous exposure to TeenFit shapes how participants understand or discuss the intervention.

All workshops are planned for a 90-minute duration and will take place in rooms equipped with whiteboards, internet access, and sufficient wall space for displaying materials. While in-person sessions will be prioritized, individual online sessions conducted via Google Meet or a similar platform will be offered when necessary. Parent and teacher workshops will be scheduled whenever possible during existing parent-teacher association meetings. Workshops will health professionals will ideally be conducted during scheduled professional development hours. If this is not possible, sessions will be held outside working hours in training rooms within the healthcare district. All workshops will involve one moderator who leads the session and two facilitators who stimulate discussion and interaction between participants while also observing and taking field notes. All sessions will be audio- and video-recorded.

### Qualitative data and data analysis

The TeenFit website will be progressively developed in collaboration with participants through an iterative and participatory process. Video recordings from the workshops will ensure that researchers who are not attending the workshop can still be involved in the co-design process. Audio recordings will be transcribed verbatim and analyzed to describe participants’ views on (1) what is needed to meet the needs of the stakeholder groups regarding the content, features, and layout of the website, and (2) barriers and facilitators to sustainable implementation of the TeenFit website in real-world settings (e.g., high schools, sports clubs, and healthcare settings).

Data collection and analysis will occur concurrently during the development phase, allowing the research team and participants to jointly interpret and refine insights as they emerge during the workshops. This back-and-forth process between ideation and visualization will guide the creation of preliminary prototypes of the TeenFit website. An initial version will be developed based on findings from the first co-creation sessions and refined through iterative feedback loops. This analytical approach is inspired by participatory action research, emphasizing the need for a thoughtful understanding of the target groups’ behaviors and context; translation of the insights gained into concrete design priorities and features, and the development of a prototype that is tested and refined [32]. Throughout the process, participants’ contributions will be systematically reviewed, discussed, and integrated with the research team’s analyses. Qualitative content analysis [33] will be used to describe the co-design process and specifically participants’ perspectives on what aspects (i.e., content, features, and layout) are important for the design of the website [33].

During the third workshop, semi-structured questions inspired by the five domains of the Consolidated Framework for Implementation Research (CFIR) [34] will be used to focus the discussion on conditions influencing the implementation of the TeenFit website. The five domains relate to the innovation: the setting, including the broader system and the specific context; the professionals; and the implementation support provided. Transcribed data from audio recordings will be analyzed using a latent reflexive thematic analysis [35]. The analysis involves inductive coding and organizing the data based on recurring concepts and patterns of shared ideas and experiences among participants, to describe and contextualize the determinants of implementation outcomes. All qualitative data analyses will be carried out using ATLAS.ti software. The results from the analysis of implementation data will be merged with results from quantitative data on the acceptability of the TeenFit website to deepen understanding of whether—and how—the identified barriers and facilitators to implementation can be addressed through website revisions and implementation support. Consequently, the second aim of this project will be addressed through a convergent mixed-methods study design [16].

### Quantitative data and analysis

Demographic data, experience during the co-design process, and acceptability of the TeenFit website will be collected as part of the quantitative measures of this project. Demographic data will be collected and include age, gender, socioeconomic status, physical activity levels, caregiver type (e.g., legal guardian, father, mother, or grandparents), teacher subject, or healthcare specialty. Socioeconomic status will be measured by asking adolescent participants the following question: “In your opinion, how is your family doing financially?” rated on a 4-point scale: 3 = *we have enough money to do anything we want*, 2 = *we have all the money we need to live a comfortable life*, 1 = *money is tight, but we are able to meet our basic needs*, and 0 = *we do not have enough money to meet basic needs*. Caregivers will be asked: “Thinking about the end of each month over the past 12 months, would you say your household generally ended up with?”: 3 = *more than enough money left over*, 2 = *some money left over*, 1 = *just enough to make ends meet*, and 0 = *not enough to make ends meet*. These questions have been evaluated and shown to provide valid approximations of family socioeconomic status [36]. Although educators and health professionals will not be asked for specific questions regarding socioeconomic status, educational attainment can serve as an indicator of socioeconomic position. Moreover, information about participants’ occupations provides an additional basis for inferring socioeconomic characteristics.

The four groups of stakeholders (i.e., adolescents, caregivers, educators, and health professionals) will also be asked about their physical activity using the Spanish version of the International Physical Activity Questionnaire – Short Form (IPAQ-SF) [37], which consists of seven items targeting vigorous and moderate physical activity as well as sedentary behavior, using open-ended 7-day recall responses.

Experience during the co-design process will be collected at the end of workshop sessions 1 and 2 using three self-developed items rated on a 5-point Likert scale, ranging from 0 (strongly disagree) to 4 (strongly agree). The items are as follows: (1) Participating in the co-design process made the TeenFit content better suited to my needs, and (2) The co-design process made me feel part of the project. The third item is tailored to adolescents, or caregivers, and health professionals. Adolescents will be asked to respond to the following statement: (3) Participating in the workshop improved my knowledge about physical activity and increased my ability to take charge of my daily physical activity. Caregivers and health professionals will be asked to respond to the following statement: Taking part in the workshop gave me new perspectives on how digital platforms can serve as tools for promoting physical activity in adolescents. These items were adapted from the instrument developed by the European IC-Health project for the Spanish cohorts Álvarez-Pérez et al. [26].

Acceptability of the TeenFit website will be collected at the end of workshop session three. Acceptability will be assessed through four open-ended questions and a 14-item questionnaire rated on a five-point Likert scale (from *strongly disagree* to *strongly agree*). The instrument was developed based on previous related studies [26, 38] and evaluates aspects such as ease of navigation, clarity of objectives and language, appropriateness of labels, and other characteristics of the TeenFit website. The specific items used for assessing acceptability are provided in **Supplemental Material 2**.

Quantitative data from these surveys will be analyzed using descriptive statistics. Means and standard deviations (SD) will be calculated for all items, and the response distribution for each item will also be analyzed. If relevant, medians and interquartile ranges (IQR) will additionally be calculated. Furthermore, results from the acceptability questionnaire will be integrated with qualitative findings from workshop 3 by presenting quantitative scores alongside qualitative summaries and illustrative quotes addressing key areas covered by the questionnaire (e.g., innovation clarity and relevance). Such a joint display will facilitate interpretation of the combined data and provide deeper insights into the integrated findings on conditions influencing the acceptability and usability of the TeenFit website[17], which are likely to affect implementation outcomes.

## ETHICS AND DISSEMINATION

This study is part of the TeenFit project: TeenFit: The Physical Activity App Finder for Adolescents. The patient and public involvement processes were approved by the Research Ethics Committee of the province of Málaga, Spain (REC ref: SICEIA-2024-003172), and the study will be conducted in accordance with the ethical principles of the Declaration of Helsinki. Participation will be voluntary, and written informed consent will be obtained at the beginning of the first workshop. For minors, consent will be obtained from both the participant and their primary caregiver.

Findings from the study, including summaries of workshop results and the final TeenFit website, will be disseminated to participants, collaborating institutions, and relevant stakeholders through reports, presentations, and online resources. Dissemination strategies will be tailored to ensure accessibility for adolescents, caregivers, teachers, and health professionals, promoting the uptake and use of the TeenFit website in real-world settings. Full results from the workshops are expected to be published in December 2027.

## DISCUSSION

This research protocol outlines a co-design and implementation approach using workshops to develop the TeenFit digital website, which promotes physical activity among adolescents. The co-design process actively involves adolescents, as well as key stakeholders such as caregivers, educators, and health professionals, ensuring that all participants can contribute as active partners throughout the development of the website. The protocol describes how participants will be involved in the co-design process, and these procedures will be iteratively refined to improve participation and ensure that the website reflects users’ needs, preferences, and experiences.

It is well known that most adolescents lead sedentary lifestyles, with lower levels of physical activity and engagement in health-promoting behaviors compared to recommended guidelines [8]. Barriers include limited knowledge of the benefits of physical activity, reduced motivation, and difficulty accessing appropriate resources [39]. Evidence-based strategies for promoting physical activity through digital interventions in this population remain limited. The TeenFit project aims to address these gaps by producing a catalog of validated physical activity apps and a user-centered website. The project will identify user needs, priorities, and preferences, as well as facilitators and barriers to adopting health-promoting behaviors through the TeenFit app finder. By involving adolescents, caregivers, educators, and health professionals in a co-design process, we believe that the website will be accessible, engaging, and tailored to users’ preferences and daily routines. This participatory approach is expected to optimize engagement, improve adherence, and enhance the reach and usability of the TeenFit website, ultimately empowering young people and their caregivers with practical, science-based tools to enhance physical activity, mental well-being, and overall health.

The TeenFit project combines a participatory co-design approach with a mixed-methods design, providing comprehensive insights into participant engagement, acceptability, and potential implementation of the website. Additionally, the use of a variety of activities during the workshops (e.g., brainstorming, real-world scenarios, creating visuals representing the group’s perceptions and experiences, and journey mapping) aligns with recommendations [40] and is likely to promote engagement and generate valuable input.

The website will include mobile apps available in the Spanish marketplace, identified through a systematic review and a parallel Play Store review, ensuring that the included apps are evidence-informed and relevant. However, the systematic and Play Store reviews are currently in the data extraction phase and have inherent limitations, including potential gaps in app availability and quality assessment. The sample size is considered sufficient for co-design and exploratory research, allowing meaningful input for website development. Although effort will be made, the study relies on a convenience sample, which may not fully represent such as populations in rural areas or reflect broader sociocultural diversity.

Overall, TeenFit will produce a validated catalog of physical activity apps, a user-centered web tool, and insights to support its implementation. Ultimately, TeenFit seeks to empower adolescents to live healthier and more active lives by offering accessible, science-based tools to enhance health and well-being.

## Data Availability

Not applicable

## Contributors

AS, MR-A, MIB-R, PH, RAL, YA-P, RER, JAB, and EM conceived the study and designed the co-design approach. MIB-R, AR-M, AA, YA-P, HC-P, and MR-A coordinated the co-design workshops with input from community and professional stakeholders. MIB-R, AR-M, AA, HC-P, YA-P, and MR-A also contributed to the development of workshop materials and data collection tools. AS, MF-L, PH, and RAL provided methodological guidance and oversaw data analysis. AS and MR-A drafted the protocol. All authors contributed to, reviewed, and approved the final version of the manuscript.

## Patient and public involvement

Patients and/or the public were involved in the design, or conduct, or reporting, or dissemination plans of this research. Refer to the Methods section for further details.

## Patient consent for publication

Not required.

## Provenance and peer review

Not commissioned; externally peer reviewed.

## Open access

This is an open access article distributed in accordance with the Creative Commons Attribution Non Commercial (CC BY--NC 4.0) license, which permits others to distribute, remix, adapt, build upon this work non--commercially, and license their derivative works on different terms, provided the original work is properly cited, appropriate credit is given, any changes made indicated, and the use is non--commercial. See: http://creativecommons.org/licenses/by-nc/4.0/.

