## Supplemental Material 1 for "A co-designed website (TeenFit) to promote physical activity in adolescents through mobile apps: a study protocol"

**TeenFit Co-Design and Implementation Sessions**

**Workshop session 1**

**Objective:** Brainstorming and real-case scenarios to create the first prototype of the TeenFit website.

**Preparation**

- Backup audio/video recorder.
- Test the availability of devices (microphones, cameras, tablets/laptops if used).
- Arrange the room for group and individual work.
- Confirm participants completed consent forms and questionnaires.
- Send a reminder email 1 day before the session.

**Introduction**

- Welcome, researchers’ introduction, session agenda, and participation rules.
- Project objective and method.
- Briefly explain physical activity habits and technology use.
- Participant introductions:
  - Adolescents: physical activity app usage and motivations.
  - Caregivers: support strategies for adolescents.
  - Professionals: guidance provided for app selection.

**Main act**

- **Part 1:** App experiences. Map experience with physical activity apps. Explore apps used, usability, and improvement points. Use colored cards/sticky notes to indicate agreement (green/yellow/red).
- **Part 2:** Explore information needs and website features. Scenario-based activities: e.g., adolescent at primary care consultation choosing an app.

**Evaluation**

- Summarize agreements and commitments.
- Explain next session: prioritization of app features and content.
- Fill out the co-design experience and sociodemographic questionnaires.

**Workshop session 2**

**Objectives:**

- Receive feedback on the first TeenFit prototype.
- Prioritize features and content for TeenFit resources.
- Identify user needs and useful information sources.

**Preparation**

- Same technical setup and room arrangement as Session 1.
- Shared whiteboards or collaborative areas for prioritization exercises.

**Introduction**

- Welcome and acknowledgment.

**Main act**

- **Part 1:** Feedback on First Prototype. Present the experience map; request feedback on content and visual representation.
- **Part 2:** Prioritization exercise. Participants review the journey map individually or in groups. Select the most important features or content. Decide which aspects require extra attention (labels, app descriptions).

**Evaluation**

- Recap agreements and priorities.
- Explain follow-up: website updates, content development, and implementation sessions (half of participants needed).
- Fill out co-design experience.

**Workshop session 3**

**Objectives:**

- Test and evaluate the real-world implementation of TeenFit.
- Collect quantitative and qualitative feedback.
- Identify barriers and facilitators to adoption.
- Assess acceptability (only in the sub-sample that did not participate in the previous workshop).

**Preparation**

- Arrange room for individual or small-group testing.
- Prepare devices with the TeenFit website preloaded.
- Ensure availability of materials (pens, paper, devices).
- Prepare surveys and observation sheets.
- Ensure technical support for troubleshooting.
- Devices with the TeenFit website preloaded.
- Surveys and observation sheets.
- Pens and paper.
- Recording equipment (with consent).

**Introduction**

- Welcome participants; overview of objectives.
- Emphasize the importance of feedback.
- **Main act Part 1:** Website interaction. Participants interact individually or in small groups. Observe interactions, challenges, and feedback.
- **Part 2:** Structured Feedback Collection. Surveys on acceptability (only in the sub-sample that did not participate in the previous workshop). Semi-structured interviews for qualitative feedback. Observation of non-verbal cues.
- **Part 3:** Discussion of barriers and facilitators for implementation in various contexts. Encourage suggestions for improvement.

**Evaluation**

- Summarize key findings.
- Explain the use of feedback for website refinement.
- Inform participants about follow-up.
- Sociodemographic questionnaires of new participants.
- Fill out the acceptability questionnaire
