## Supplemental Material 2 for "A co-designed website (TeenFit) to promote physical activity in adolescents through mobile apps: a study protocol"

**Adaptation of the acceptability questionnaire:**

1. **What do you think about the title of the website?**
   (Open-ended question)
2. **The language used on the website was easy to understand.**

- Strongly disagree
- Disagree

Neither agree nor disagree

Agree

- Strongly Agree

2. **It was easy to navigate through the website and to choose the app I was looking for.**

- Strongly disagree
- Disagree

Neither agree nor disagree

- Agree
- Strongly Agree

3. **The information presented was organized coherently and clearly.**

- Strongly disagree
- Disagree

Neither agree nor disagree

- Agree
- Strongly agree

4. **The overall quality of the website design and its content seemed to me:**

- Strongly disagree
- Disagree

Neither agree nor disagree

- Agree
- Strongly agree

5. **The website objectives were clear.**

- Strongly disagree
- Disagree

Neither agree nor disagree

- Agree
- Strongly agree

6. **The website content was consistent with the stated objectives and with the intended audience.**

- Strongly disagree
- Disagree

Neither agree nor disagree

- Agree
- Strongly agree

7. **I found the website interesting.**

- Strongly disagree
- Disagree

Neither agree nor disagree

- Agree
- Strongly agree

8. **This website met my expectations.**

- Strongly disagree
- Disagree

Neither agree nor disagree

- Agree
- Strongly agree

9. **The description of each app and its features was clear and easy to understand, avoiding possible misinterpretations.**

- Strongly disagree
- Disagree

Neither agree nor disagree

- Agree
- Strongly agree

10. **The titles, buttons, and links on the website clearly matched the content or pages they led to.**

- Strongly disagree
- Disagree
- Undecided

Neither agree nor disagree

Agree

- Strongly agree

11. **The information and recommendations on the website were useful for helping me understand how to choose and use physical activity apps.**

- Strongly disagree
- Disagree

Neither agree nor disagree

- Agree
- Strongly agree

12. **The tools or features provided (e.g., filters, app descriptions, ratings) helped me evaluate which apps were most suitable for me.**

- Strongly disagree
- Disagree

Neither agree nor disagree

- Agree
- Strongly agree

13. **Do you think this website has helped you improve your ability to practice more physical activity by learning how to find relevant and reliable physical activity apps in the marketplace?**

- Strongly disagree
- Disagree

Neither agree nor disagree

- Agree
- Strongly agree

14. **I would recommend this website to other people.**

- Strongly disagree
- Disagree

Neither agree nor disagree

- Agree
- Strongly agree

15. **The amount of time required to find the physical activity app that suits you was:**

- Appropriate
- Excessive, required too much time
- Insufficient, required too little time

16. **Please provide a summary of the website’s strengths and weaknesses.**
    (Open-ended question)
17. **Please provide brief suggestions on how to improve the website.**
    (Open-ended question)
18. **What are the main points you have learned throughout this website?**
    (Open-ended question)
